# Comparison of nasopharyngeal swab vs. lower respiratory tract specimen PCR for the diagnosis of *Pneumocystis jirovecii* pneumonia

**DOI:** 10.1101/2023.10.28.23297710

**Authors:** Rusheng Chew, Sarah Tozer, Kimberly Ulett, David L. Paterson, David Whiley, Theo Sloots, David Fielding, Christopher Zappala, Farzad Bashirzadeh, Justin Hundloe, Cheryl Bletchley, Marion L. Woods

**Author notes:** **Corresponding author details** Dr Rusheng Chew, Address: Mahidol Oxford Tropical Medicine Research Unit, c/o Faculty of Tropical Medicine, Mahidol University, 3^rd^ floor, 60^th^ Anniversary Chalermprakiat Building, 420/6 Ratchawithi Road, Ratchathewi, Bangkok 10400, Thailand.

## Abstract

**Background and objective:** Diagnosis of *P. jirovecii* pneumonia (PJP) is by PCR on lower respiratory tract specimens, the collection of which is not always well-tolerated and requires trained staff and costly equipment not usually available in low-resource settings. We aimed to evaluate *P. jirovecii* PCR performed on nasopharyngeal swabs (NPS) as a diagnostic test for PJP, as well as the impact of specimen quality on test performance.

**Methods:** Patients with clinically-suspected PJP in public hospitals in Queensland, Australia, who had quantitative *P. jirovecii* PCR performed on lower respiratory tract specimens from 1 January 2015 to 31 December 2016, and also had NPS collected by healthcare staff within seven days of lower respiratory tract specimen collection were included in this retrospective cohort study. Quantitative *P. jirovecii* PCR was performed, and sensitivity, specificity, and positive and negative predictive values were calculated. Specimen quality was assessed by quantifying endogenous retrovirus 3 (ERV3) loads, with higher values indicating better specimen quality.

**Results:** One hundred and eleven patients were included. The sensitivity of NPS *P. jirovecii* PCR was 0.66 and specificity was 1.0. The positive predictive value was 1.0 and the negative predictive value was 0.63. Median ERV3 loads in lower respiratory tract specimens and NPS were not significantly different in true positive vs. true negative patients, but was significantly higher in true positives vs. false negatives (7.55×10^2^ vs. 3.67×10^2^; *P*=0.05).

**Conclusion:** *P. jirovecii* PCR on NPS was highly specific but poorly sensitive. Proper specimen collection is essential to ensure adequate quality and prevent misclassification.

**Summary at a Glance:** Using nasopharyngeal swabs instead of lower respiratory tract specimens for PCR to diagnose *P. jirovecii* pneumonia (PJP) may be better tolerated and improve diagnostic accessibility. In this two-year retrospective cohort study of patients with clinically-suspected PJP from Queensland, Australia, *P. jirovecii* PCR on NPS had high specificity but low sensitivity.

## Introduction

The unicellular fungus *Pneumocystis jirovecii* is an opportunistic pathogen causing pneumonia in immunocompromised hosts,^1^ such as those with HIV/AIDS.^2^ It is also the commonest fungal cause of pneumonia in non-HIV infected children under 5 years.^3^ Despite global improvements in HIV prevention and treatment, the burden of *P. jirovecii* pneumonia (PJP) remains considerable due to the increasing use of immunosuppressive agents for transplantation, malignancy, and autoimmune disorders, along with sub-optimal control of the HIV/AIDS epidemic in developing countries.^4, 5^

Because *P. jirovecii* is not culturable in vitro, polymerase chain reaction (PCR) is the gold standard diagnostic method in many countries.^6, 7^ *P. jirovecii* PCR is generally performed on lower respiratory tract specimens, such as induced sputum or broncho-alveolar lavage fluid. However, patients may be too frail, young, or hypoxic to undergo such procedures, especially fibre-optic bronchoscopy which may result in unintended morbidity and mortality.^8^ Lower respiratory tract specimen collection may be invasive and carries a risk of patient discomfort, such as bronchospasm from sputum induction.^9^ It also requires trained staff as well as costly equipment and facilities, such as negative pressure rooms, resulting in service inequity for patients in rural or remote locations and developing countries.^7, 8^

There is, therefore, a need for specimens obtainable via low-cost, minimally invasive methods on which *P. jirovecii* PCR can be performed, but which do not compromise test performance. Nasopharyngeal swabs may be such an alternative. Nevertheless, to our knowledge, the only evidence comparing nasopharyngeal and lower respiratory tract specimens comes from children in a developing country high HIV prevalence setting published over a decade ago,^10^ indicating a gap in the contemporary evidence. In this study, we aimed to evaluate *P. jirovecii* PCR performed on nasopharyngeal swabs as a diagnostic test for PJP, as well as the impact of specimen quality on test performance.

## Methods

### Study design

Retrospective cohort study. The study STROBE checklist can be found in Supporting Document S1.

### Setting and participants

Queensland is an Australian state with an estimated HIV prevalence of 0.14%.^11^ Eligible patients were those with clinically suspected PJP in public sector hospitals who had *P. jirovecii* PCR performed on lower respiratory tract specimens (either induced sputum or broncho-alveolar lavage fluid) over two years from 1 January 2015 to 31 December 2016, and also had nasopharyngeal swabs collected by healthcare staff within seven days of lower respiratory tract specimen collection. Patients were identified through a state-wide computerized database (AUSLAB; Citadel Health, Canberra, Australia), which contains records of all laboratory investigations requested in public sector healthcare facilities.

### PCR method

*P. jirovecii* PCR was performed on lower respiratory tract specimens and nasopharyngeal swabs on two PCR platforms; a positive result was returned if either detected *P. jirovecii* DNA.

The PCR detection and subsequent quantification of *P*.*jirovecii* was performed using an in-house real-time method on a Rotor-Gene instrument (QIAGEN NV, Venlo, The Netherlands). The mix was prepared using the QIAGEN QuantiTect PCR kit (QIAGEN NV, Venlo, The Netherlands) incorporating specific primers and probe targeting the 5s ribosomal RNA gene, details of which are as follows: forward primer (PCP-TM-5s-F) AGTTACGGCGATACCTCAGAGAATATAC; reverse primer (PCP-TM-5s-R) GCTACAGCACGTCGTATTCCCATA; probe (PCP-TM-5s-probe) FAM-TCACCCACTATAGTACTGACGACGCCCTT-BHQ.

The control and quantitative standards were prepared in-house using extracted patient material with high copy numbers. The standard for the qualitative assay was prepared at 2.5×10^10^ copies/ml, while four standards were prepared to provide a standard curve with the range 8.0×10^4^ to 8.0×10^7^ copies/ml for the quantitative assay. The standard curve was plotted with the supplied Rotor-Gene software using the quantitation option, and quantitative results for samples were determined from this plot by the software. Results were expressed as copies/ml.

### Assessment of specimen quality

Specimen quality was assessed through human DNA quantification using endogenous retrovirus 3 (ERV3) as a surrogate marker, per the method of Alsaleh et al.^12^ Because each diploid human cell contains two copies of this retrovirus, ERV3 PCR allows accurate quantification of the number of human cells present in a sample; the higher the ERV3 load, the better the quality of the specimen. Its use for this purpose is well-established, with high analytical sensitivity and specificity.^13^

### Statistical analysis

Patients for whom nasopharyngeal swab *P. jirovecii* PCR results were unavailable were excluded from the analysis. Of the remaining patients, only those with non-missing values of ERV3 load were included in the assessment of specimen quality. Patients who tested negative for ERV3 i.e., had an ERV3 load of zero, were not considered to have missing values for ERV3 load.

The diagnostic sensitivity and specificity, as well as the positive and negative predictive values (PPV and NPV), of nasopharyngeal swabs for PJP were calculated.

To assess the impact of specimen quality, ERV3 loads were compared as follows: in lower respiratory tract specimens in patients with and without PJP (i.e., true positives and true negatives); in nasopharyngeal swab specimens from true positives and true negatives; and in nasopharyngeal swabs from true positives and false negatives, using Mann-Whitney U tests.

Analyses were performed using Stata 17 (StataCorp, College Station, USA). *P* values ≤0.05 were considered significant. Raw data are available upon request from the corresponding author.

### Ethical approval

Ethical approval was obtained from the Royal Brisbane and Women’s Hospital Ethics Committee (HREC/14/QRBW/19).

### Funding

This research was funded by a Study, Education, and Research Trust Fund grant from Pathology Queensland. RC was supported by the UK Government through a Commonwealth Scholarship and the Royal Australasian College of Physicians through the Bushell Travelling Fellowship in Medicine or the Allied Sciences. The funders had no role in study design, data collection, data analysis, data interpretation, or writing of the manuscript.

## Results

One hundred and eleven patients met the inclusion criteria; their characteristics are summarized in Table 1. Data on ERV3 loads in lower respiratory tract specimens and nasopharyngeal swabs were missing in 3/111 (2.7%) and 13/111 (11.7%) patients, respectively.

**Table 1.** Participant characteristics. Patients with *Pneumocystis jirovecii* pneumonia (PJP) were those who were clinically suspected of having the disease and had positive *P. jirovecii* polymerase chain reaction (PCR) results on lower respiratory tract specimens (induced sputum or broncho-alveolar lavage fluid). HIV, human immunodeficiency virus; IQR, interquartile range.

|  | Patients without PJP<br>(N=40) | Patients with PJP<br>(N=71) |
| --- | --- | --- |
| **Age (years)** |  |  |
| Median (IQR) | 62 (46 – 73) | 66 (55 – 72) |
| **Sex (n, %)** |  |  |
| Male | 19 (17.1) | 32 (28.8) |
| Female | 21 (18.9) | 39 (35.1) |
| **Immunosuppression type (n, %)** |  |  |
| Haematological malignancy or transplant | 14 (12.6) | 13 (11.7) |
| Solid organ malignancy | 7 (6.3) | 28 (25.2) |
| Solid organ transplant | 7 (6.3) | 3 (2.7) |
| HIV-positive | 1 (0.9) | 11 (9.9) |
| Other iatrogenic immunosuppression | 4 (3.6) | 14 (12.6) |
| Other non-iatrogenic immunosuppression | 7 (6.3) | 2 (1.8) |
| **Nasopharyngeal swab *P. jirovecii* PCR result (n, %)** |  |  |
| Not detected | 40 (36.0) | 24 (21.6) |
| Detected | 0 (0.0) | 47 (42.3) |

From the data in Table 1, the diagnostic sensitivity of nasopharyngeal swab *P. jirovecii* PCR was 0.66, while its specificity was 1.0. The positive predictive value was 1.0 and the negative predictive value was 0.63.

Median ERV3 loads in lower respiratory tract specimens were not significantly different in true positive and true negative patients (4.25×10^3^ vs. 8.56×10^3^; *P*=0.06). Median nasopharyngeal swab ERV3 loads were also not significantly different between these two groups (6.20×10^2^ vs. 1.71×10^3^; *P*=0.07). However, the median ERV3 load in nasopharyngeal swabs from true positives was significantly higher than in false negatives (7.55×10^2^ vs. 3.67×10^2^; *P*=0.05).

## Discussion

We found nasopharyngeal swabs for diagnosis of *P. jirovecii* PCR to be highly specific but poorly sensitive for diagnosing PJP compared to lower respiratory tract specimens. Given the absence of false positives, therefore, this indicates that a positive PCR on nasopharyngeal swab is sufficient to diagnose PJP. It follows also that our results suggest that colonization of the upper respiratory tract by *P. jirovecii* is not a barrier to performing PCR on nasopharyngeal swabs for this purpose in adults.

Additionally, we showed that false negative nasopharyngeal swabs contained significantly lower ERV3 loads than true positives, demonstrating the importance of proper specimen collection to ensure adequate quality and prevent misclassification.^14^ One method of improving the chances of adequate specimen collection is to collect more than one swab for testing in parallel. This has the added benefit of increasing sensitivity, since the sensitivity would be expected to increase to 88% with two swabs and to 96% with three. Such augmentation of the performance of low-sensitivity assays with multiple testing has been demonstrated with SARS-CoV-2 rapid antigen tests in a real-world population during the current COVID-19 pandemic.^15^

To our knowledge, this is the first study of the use of *P. jirovecii* PCR on nasopharyngeal swabs for PJP diagnosis in immunosuppressed adult patients living in a developed country, low HIV prevalence setting. Another benefit of using nasopharyngeal swabs is the ability to test for concurrent or alternative diagnoses simultaneously e.g., with a multiplex PCR panel; in our cohort, 12/71 patients with PJP and 6/41 without PJP also had a PCR-confirmed viral respiratory infection.

Our results are not generalizable to children, as none featured in our dataset. The number of patients in this study was small which, together with its observational nature focused on patients with high pre-test probabilities for PJP, are potential sources of bias. This risk was mitigated by including patients fitting the inclusion criteria from all Queensland public hospitals over the span of two years. While we expect *P. jirovecii* PCR on nasopharyngeal swabs to perform similarly in adult patients from other low HIV prevalence settings, this hypothesis should be tested as part of future research. Ideally this would be a prospective study including all patients clinically suspected of having PJP in a wider range of settings and patient demographics, with paired lower respiratory tract specimens and two to three NP swabs collected from each patient using the correct technique on the same day.

In conclusion, our hypothesis-generating study demonstrates the potential utility of NP swabs for the diagnosis of PJP and lends support for a large-scale prospective study to address this important question, the answer to which will have immediate clinical impact.

## Data availability

The data that support the findings of this study are available on request from the corresponding author. The data are not publicly available due to privacy or ethical restrictions.

## References

1 Sokulska M, Kicia M, Wesołowska M, Hendrich AB. Pneumocystis jirovecii—from a commensal to pathogen: clinical and diagnostic review. Parasitol Res. 2015; 114: 3577–85.

2 Morris A, Lundgren JD, Masur H, Walzer PD, Hanson DL, Frederick T, et al. Current epidemiology of Pneumocystis pneumonia. Emerg Infect Dis. 2004; 10: 1713–20.

3 3 Pneumonia Etiology Research for Child Health (PERCH) Study Group. Causes of severe pneumonia requiring hospital admission in children without HIV infection from Africa and Asia: the PERCH multi-country case-control study. Lancet. 2019; 394: 757–79.

4 Cillóniz C, Dominedò C, Álvarez-Martínez MJ, Moreno A, García F, Torres A, et al. Pneumocystis pneumonia in the twenty-first century: HIV-infected versus HIV-uninfected patients. Expert Rev Anti Infect Ther. 2019; 17: 787–801.

5 Pereira-Díaz E, Moreno-Verdejo F, De la Horra C, Guerrero JA, Calderón EJ, Medrano FJ. Changing Trends in the Epidemiology and Risk Factors of Pneumocystis Pneumonia in Spain. Front Public Health. 2019; 7: 275.

6 Cushion MT, Tisdale-Macioce N, Sayson SG, Porollo A. The Persistent Challenge of Pneumocystis Growth Outside the Mammalian Lung: Past and Future Approaches. Front Microbiol. 2021; 12: 681474.

7 Bateman M, Oladele R, Kolls JK. Diagnosing Pneumocystis jirovecii pneumonia: A review of current methods and novel approaches. Med Mycol. 2020; 58: 1015–28.

8 Du Rand IA, Blaikley J, Booton R, Chaudhuri N, Gupta V, Khalid S, et al. British Thoracic Society guideline for diagnostic flexible bronchoscopy in adults: accredited by NICE. Thorax. 2013; 68 Suppl 1: i1–i44.

9 Choe PG, Kang YM, Kim G, Park WB, Park SW, Kim HB, et al. Diagnostic value of direct fluorescence antibody staining for detecting Pneumocystis jirovecii in expectorated sputum from patients with HIV infection. Med Mycol. 2014; 52: 326–30.

10 Samuel CM, Whitelaw A, Corcoran C, Morrow B, Hsiao NY, Zampoli M, et al. Improved detection of Pneumocystis jirovecii in upper and lower respiratory tract specimens from children with suspected pneumocystis pneumonia using real-time PCR: a prospective study. BMC Infect Dis. 2011; 11: 329.

11 State of Queensland (Queensland Health). 2020. HIV in Queensland 2018. https://www.health.qld.gov.au/__data/assets/pdf_file/0025/940237/hiv-in-queensland-2018.pdf. Accessed: 23 February 2022.

12 Alsaleh AN, Whiley DM, Bialasiewicz S, Lambert SB, Ware RS, Nissen MD, et al. Nasal swab samples and real-time polymerase chain reaction assays in community-based, longitudinal studies of respiratory viruses: the importance of sample integrity and quality control. BMC Infect Dis. 2014; 14: 15.

13 Yuan CC, Miley W, Waters D. A quantification of human cells using an ERV-3 real time PCR assay. J Virol Methods. 2001; 91: 109–17.

14 Marty FM, Chen K, Verrill KA. How to Obtain a Nasopharyngeal Swab Specimen. N Engl J Med. 2020; 382: e76.

15 Dřevínek P, Hurych J, Kepka Z, et al. The sensitivity of SARS-CoV-2 antigen tests in the view of large-scale testing. Epidemiol Mikrobiol Imunol. 2021; 70: 156–60.

